# Psychological, social and financial impact of COVID-19 on culturally and linguistically diverse communities: a cross-sectional Australian study

**DOI:** 10.1101/2021.10.19.21265230

**Authors:** DM Muscat, J Ayre, O Mac, C Batcup, E Cvejic, K Pickles, H Dolan, C Bonner, D Mouwad, D Zachariah, U Turalic, Y Santalucia, T Chen, G Vasic, KJ McCaffery

## Abstract

**Objective:** This study aimed to explore the psychological, social, and financial impacts of COVID-19 on culturally and linguistically diverse communities in Australia.

**Design:** Cross-sectional survey informed by the Framework for Culturally Competent Health Research conducted between March and July, 2021.

**Setting:** Participants were recruited from Greater Western Sydney, New South Wales, Australia.

**Participants:** 708 community members who speak a language other than English at home participated (mean age: 45.4years [range 18–91]; 88% [n=622] born outside of Australia).

**Outcome measures:** Fifteen items regarding impacts of COVID-19, adapted from validated scales, previous surveys or co-designed in partnership with Multicultural Health and interpreter service staff. Logistic regression models (using post-stratification weighted frequencies) identified factors associated with psychological, social, and financial impacts. Surveys were available in English or translated (11 languages).

**Results:** Even prior to the COVID-19 outbreak in Sydney, 25% of the sample reported feeling nervous or stressed most/all of the time and 22% felt lonely or alone most/all of the time. One quarter of participants reported negative impacts on their spousal relationships as a result of COVID-19 and most parents reported that their children were less active (64%), had more screen time (63%), and were finding school harder (45%). Mean financial burden was 2.9/5 (95%CI=2.8 to 2.9). Regression analyses consistently showed distinct impact patterns for different language groups and more negative outcomes for those with comorbidities.

**Conclusion:** Culturally and linguistically diverse communities experience significant psychological, social and financial impacts of COVID-19, with distinct impact patterns across language groups. A whole-of-government approach with policy and sustainable infrastructure is needed to co-design innovative, tailored and culturally-safe COVID-19 support packages.

**ARTICLE SUMMARY:** **Strengths and limitations of this study**

- This is the largest Australian survey exploring the impacts of COVID-19 among people who primarily speak a language other than English, enabled through recruitment methods that are inclusive and reduce barriers to participation (e.g. translated surveys, engagement of interpreters and multicultural health staff who are trusted in their communities, and use of multiple recruitment methods including through community events and networks).
- This study was co-designed by researchers and multicultural health service staff, in alignment with the Framework of Culturally Competent Health Research.
- To reduce survey length and burden on participants we purposefully selected a small number of items from validated measures or our previous research to explore psychological, social and financial impacts, or co-designed them specifically for this study.
- We used convenience sampling methods and self-report may have introduced recall and social desirability bias.
- We are unable to explore changes in impacts of COVID-19 over time.

## INTRODUCTION

The COVID-19 pandemic has not impacted all populations equally. Ethnic minority groups in countries across the globe have been disproportionately affected, with higher rates of infection, greater risk of morbidity, higher critical care admissions and mortality, and poorer mental health and financial outcomes (1-6). Such differences reflect pre-existing health disparities and underlying social, economic and political inequalities; ethnic minority groups experience a higher prevalence of comorbidities associated with poor COVID-19 outcomes (e.g. cardiovascular conditions), greater social deprivation and differences in occupational and environmental risk (7-9). The additional burden of structural racism also impacts care seeking and quality of care (7).

While the data tells a clear story of cultural disadvantage in the United States, Canada, the United Kingdom and several Nordic countries, there remains limited evidence of the impact of COVID-19 on culturally and linguistically diverse groups in Australia despite being one of the most culturally diverse nations worldwide. Nationally representative surveys exploring the financial, social and psychological impacts of the pandemic (see, for example, (10)) have not investigated culturally and linguistically diverse populations, and there remains a lack of disaggregated data related to COVID-19. A similar trend is observed worldwide (11).

Research to date (both in Australia and internationally) has also been limited in its engagement and co-production with diverse communities. This has been exacerbated by online recruitment methods (e.g. via social media networks or market research companies) and English-language data collection, which tends to exclude those who speak a language other than English as their primary language.

The few studies which have been conducted have highlighted important impacts of the pandemic for our diverse communities (12, 13). In a study of 656 refugees and asylum seekers from Arabic, Farsi, Tamil or English-speaking backgrounds who had arrived in Australia within the last 10 years, approximately one in five participants reported experiencing employment loss or decline due to COVID-19, with prevalent stressors related to COVID-19 infection including worries about being infected (66.5%), of a loved one being infected (72.1%) or infecting others (47.7%) (14). Social stressors as a consequence of the pandemic were also common, including school closures (46.7%), reduced social activities (46.6%), and having to remain at home (41.3%), and these stressors predicted increased depression symptoms and disability outcomes (14). However, the experiences of refugees and asylum seekers are unique and may not reflect the experience of migrants or those who speak a language other than English at home who have not been forced to flee their home country. Both perspectives are critically important and necessary to provide a complete picture of impacts of COVID-19 on culturally and linguistically diverse groups in Australia.

Our own Australian surveys (and others – see, (12, 13)) have shown some differences in financial and psychological impacts of COVID-19 those for who spoke a language other than English at home compared to those for whom English is their primary language. A survey of 4362 Australians conducted in April 2020, for example, showed that participants who spoke a language other than English at home rated the financial impact of COVID-19 as higher, were more likely to feel nervous or stressed as a result of the pandemic compared with those who primarily spoke English at home (15) and had greater anxiety. However, 75% of participants in this survey were born in Australia and only 274 (6%) reported that they did not speak English as their main language at home. As such, our previous findings are limited in their ability to inform appropriate and tailored support for Australian communities that are typically understudied and underserved, such as those from different cultural and language groups.

The aims of this study were to:

1. Explore the psychological, social, and financial impact of the COVID-19 pandemic on culturally and linguistically diverse communities in Greater Western Sydney in New South Wales (NSW), Australia.
2. Examine demographic factors associated with these impacts.

## METHODS

### Study design

This study involved a cross-sectional survey with 11 language groups, approved by Western Sydney Local Health District Human Research Ethics Committee (Project number 2020/ETH03085)

### Patient and public involvement

This study was co-designed by researchers, bilingual community members and Multicultural Health and Health Care Interpreter Service staff, and informed by the Framework for Culturally Competent Health Research (16).

### Setting

The survey was conducted from 21 March to 9 July, 2021. During this period, rollout of the COVID-19 vaccines had begun across Australia, and daily cases in New South Wales (NSW) were very low by international standards, ranging from 0 – 46 positive cases from a population of approximately 8 million people (17). A ‘stay at home’ order across Greater Sydney due to rising cases began on June 23^rd^ (18). On the day the survey closed the NSW daily case count was 45, and 24% of the population had received one COVID-19 vaccination (19).

Participants were recruited from Greater Western Sydney in New South Wales, Australia from three adjoining regions with high cultural diversity: Western Sydney, South Western Sydney, and Nepean Blue Mountains. Up to 39% of residents in these regions were born overseas in non-English speaking countries (20).

### Participants

Participants were eligible to take part if they were aged 18 years or over and spoke one of the following as their main language at home: Arabic, Assyrian, Chinese, Croatian, Dari, Dinka, Hindi, Khmer, Samoan, Tongan, Spanish. Through iterative discussions with Multicultural Health and Health Care Interpreter Service staff in each participating Local Health District, we selected eleven language groups that would provide broad coverage across different global regions, and groups with varying average levels of English language proficiency (based on 2016 Australian census data; (21)), varying access to translated materials, and varying degrees of reading skill in their main language spoken at home.

### Recruitment

Participants were recruited through bilingual Multicultural Health staff and Health Care Interpreter Service staff. Multicultural Health staff recruited participants through their existing networks, community events and community champions. Health Care Interpreter Service staff recruited participants at the end of a medical appointment and via their community network. The survey was hosted online using the web-based survey platform Qualtrics. Potential participants were offered two means of taking part: completing the survey themselves online (available in English or translated), or with assistance from bilingual staff or an interpreter. To ensure consistency in the phrases used for assisted survey completion, translated versions of the survey were provided to all staff assisting with survey completion. Translations were completed by translators with National Accreditation Authority for Translators and Interpreters (NAATI) accreditation where possible.

### Measures

Demographic survey items relevant to this study included age, gender, education, whether born in Australia, years living in Australia, main language spoken at home, self-reported English language proficiency, chronic disease status, and a single-item health literacy screener (22). The socioeconomic status of the area of residence for each individual was defined based on the SEIFA Index of Relative Socioeconomic Advantage and Disadvantage (IRSAD (23)). IRSAD aligns the statistical local area with a decile ranking (1–10), with lower scores indicating greater socioeconomic disadvantage. The IRSAD decile was not available for some participants (n=5), for example, because they had entered digits that did not correspond to a valid Australian postcode. IRSAD decile for these participants was replaced with the median IRSAD decile for speakers of the same language in the sample. For the analysis, IRSAD deciles were recoded into quintiles, and dichotomised (lowest quintile vs other).

Fifteen items regarding the impacts of COVID-19 were developed for this survey study. See Table 1. Items related to the psychological and financial impacts were adapted from validated scales (24) and/or our previous work (15). Questions regarding social impacts (including impacts on relationships and children) were co-designed in partnership with Multicultural Health and Health Care Interpreter Service staff. Items had fixed yes/no and Likert-type responses. Items were translated into 11 languages.

**Table 1.**
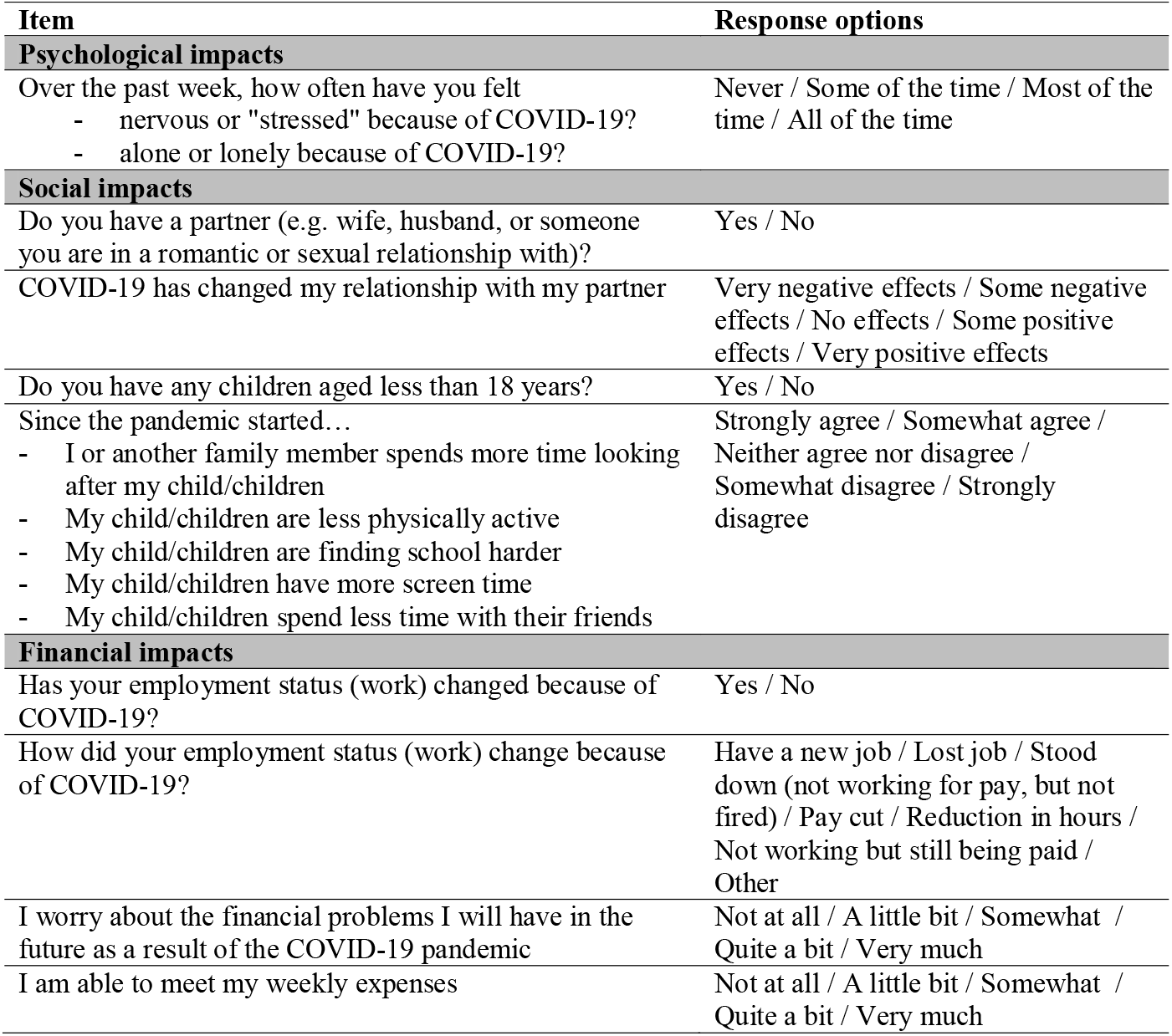
Survey items, including response options.

### Analysis

Quantitative data were analysed using IBM SPSS Statistics Version 24. Descriptive statistics were generated for demographic characteristics of the analysed sample. Frequencies were weighted (using post-stratification weighting) to reflect each language group’s gender and age group distribution (18-29 years, 30-49 years, 50-69 years, ≥70 years) based on 2016 census data for Western Sydney, South Western Sydney, and Nepean Blue Mountains’ combined populations (21). All frequencies presented in the results section are weighted. A single participant indicated their gender as ‘other’ and was unable to be included in weighted analyses. Total recruitment for the Spanish language group was low (<50), with notable gaps for some age groups. For this reason, results for this language group are not presented in the statistical analyses, but are included in total frequencies.

Survey items about psychological, financial and relationship impacts were re-coded to reflect the categories presented in Tables 3 and 4, to facilitate a more meaningful interpretation of the results. A mean ‘perceived financial burden’ score was also calculated by averaging the two questions about financial impacts: a) worry about financial problems and b) ability to meet weekly expenses. Higher scores indicate greater perceived financial burden. Similarly, a mean score for the impact on children was calculated by averaging questions related to four impacts: physical activity, screen time, schooling and time with friends. Higher scores indicate more negative impacts on children.

Multivariable linear regression models were used to analyse perceived financial burden (averaged across two impacts) and impacts on children (averaged across four impacts). Binomial logistic regression models were used to analyse psychological impacts (feeling lonely or alone; feeling nervous or stressed) and impact on relationships. Age group, gender, chronic illness, education, health literacy, English-language proficiency, years lived in Australia, language group and IRSAD quintile were included in each model. Regression models predicting impacts on relationships also controlled for perceived public health threat of COVID-19, perceived financial burden and psychological variables; models predicting psychological impacts controlled for perceived public health threat of COVID-19 and perceived financial burden. All regression models also controlled for whether participants completed the survey before or after 23^rd^ June, when restrictions were announced for all of Greater Sydney (18).

## RESULTS

### Sample characteristics

We had a total of 708 respondents (442 [62.4%] self-completed, 266 [37.6%] received assistance through an interpreter). Sample characteristics are summarised in Table 2. The mean age was 45.4 years (standard error [SE] 0.78; range 18–91 years), and 51% of respondents were female (n=363). Most participants (88%, n=622) were born in a country other than Australia; 31% reported that they did not speak English well or at all (n=220); 70% had no tertiary qualifications (n=497). Inadequate health literacy was identified for 59% of the sample (n=290).

**Table 2.** Descriptive statistics of analysed sample (N=708).

| **Variable** | **N** | **%** |
| --- | --- | --- |
| **Age group** |  |  |
| 18-29 | 147 | 20.7 |
| 30-49 | 295 | 41.8 |
| 50-69 | 193 | 27.3 |
| >70 | 72 | 10.2 |
| **Gender\*** |  |  |
| Male | 344 | 48.6 |
| Female | 363 | 51.4 |
| **Language** |  |  |
| Assyrian | 133 | 18.8 |
| Croatian | 121 | 6.2 |
| Arabic | 80 | 11.3 |
| Chinese | 76 | 10.7 |
| Dinka | 63 | 8.9 |
| Khmer | 63 | 8.9 |
| Dari | 44 | 6.2 |
| Spanish** | 43 | 6.1 |
| Hindi | 42 | 5.9 |
| Samoan/Tongan | 42 | 5.9 |
| **English language proficiency (How well do you speak English?)** |  |  |
| Very well/ well | 487 | 68.9 |
| Not well/not at all | 220 | 31.1 |
| **Literacy in a language other than English (How well do you read in your main language?)** |  |  |
| Very well/ well | 589 | 83.4 |
| Not well/not at all | 118 | 16.6 |
| **Health literacy\*\*\*** |  |  |
| Adequate | 417 | 58.9 |
| Inadequate | 290 | 41.1 |
| **Highest level of education** |  |  |
| Less than year 12(less than high school) | 115 | 16.2 |
| Year 12 (high school graduate) | 133 | 18.9 |
| Certificate level I to IV / Advanced diploma and diploma level | 249 | 35.3 |
| Bachelor degree level and above | 210 | 29.7 |
| **Years living in Australia** |  |  |
| 5 years or less | 120 | 16.9 |
| 6 to 10 years | 104 | 14.7 |
| More than 10 years | 398 | 56.4 |
| Born in Australia | 85 | 12.0 |
| **IRSAD quintile** |  |  |
| 1 (Lowest) | 224 | 31.7 |
| 2 | 140 | 19.8 |
| 3 | 125 | 17.7 |
| 4 | 140 | 19.8 |
| 5 (Highest) | 87 | 12.3 |
| **Total** | **707** |  |

NB: Frequencies are weighted (using post-stratification weighting) to reflect each language group's gender and age group distribution (18-29 years, 30-49 years, 50-69 years, ≥70 years) based on 2016 census data for Western Sydney, South Western Sydney, and Nepean Blue Mountains' combined populations (21).
\* 1 respondent indicated 'other/prefer not to say'
\*\* Spanish language group had substantial gaps in recruitment across age groups;
\*\*\* Based on the Single Item Literacy Screener (SILS) (22).

### Psychological impacts

Overall, 25.3% of participants reported feeling nervous or stressed most or all of the time over the past week. The oldest age group (70 years or more) had the highest proportion of participants reporting feeling nervous or stressed most or all of the time (35.0%, n=25) while the youngest age group (30 years or below) had the lowest proportion (20.9%, n=31). A higher proportion of females reported increased nervousness or stress (29.0%; n=105) compared to males (21.3%; n=73). 30.7% (n=89) of participants with inadequate health literacy and 21.4% (n=89) of participants with adequate health literacy reported feeling nervous or stressed most or all of the time. In terms of language groups, this ranged from 6% (n=5) for Chinese speakers to 38% (n=24) for Dinka speakers. See Tables 3 and 4. In the multivariable regression model when sociodemographic factors were controlled for, female gender (p=0.04), having two or more chronic illnesses (p<0.001) and language group (p<0.001) remained significantly associated with increased nervousness or stress, as did higher perceived financial burden (p<0.001). See Supplementary Table 1.

**Table 3.** Psychological, social and financial impacts by gender, age group, health literacy, IRSAD quintile and number of comorbidities (n=707)*

|  | Gender |  | Age group |  |  |  | Health literacy |  | IRSAD quintile |  | Comorbidities** |  |  |
| --- | --- | --- | --- | --- | --- | --- | --- | --- | --- | --- | --- | --- | --- |
|  | Male | Female | <30 | 30-49 | 50-69 | 70+** | Inadequate | Adequate | Lowest | Not lowest | 0 | 1 | 2 + |
|  | n (%) | n (%) | n (%) | n (%) | n (%) | n (%) | n (%) | n (%) | n (%) | n (%) | n (%) | n (%) | n (%) |
| **<u>Psychological impacts</u>** |  |  |  |  |  |  |  |  |  |  |  |  |  |
| Nervous or stressed | 73 (21.3) | 105 (29.0) | 31 (20.9) | 65 (22.0) | 58 (29.9) | 25 (35.0) | 89 (30.7) | 89 (21.4) | 61 (27.2) | 118 (24.4) | 85 (20.1) | 46 (29.5) | 48 (36.6) |
| Alone or lonely | 75 (21.8) | 83 (22.8) | 30 (20.2) | 52 (17.6) | 44 (22.6) | 33 (45.5) | 81 (27.8) | 77 (18.5) | 44 (19.7) | 114 (23.5) | 73 (17.4) | 37 (23.7) | 48 (36.5) |
| **<u>Social impacts\*\*\*</u>** |  |  |  |  |  |  |  |  |  |  |  |  |  |
| Negative impact on relationship | 49 (23.7) | 53 (27.3) | 23 (47.7) | 39 (19.7) | 32 (27.2) | 8 (21.6) | 49 (27.8) | 53 (23.6) | 14 (12.8) | 88 (30.3) | 54 (23.9) | 23 (25.8) | 24 (29.5) |
| More time looking after children | 99 (77.2) | 92 (68.6) | 8 (64.8) | 148 (74.1) | 34 (70.2) | - | 76 (75.7) | 114 (71.0) | 61 (72.8) | 130 (72.8) | 128 (69.9) | 41 (84.9) | 22 (71.4) |
| More screen time | 85 (66.4) | 81 (60.2) | 6 (46.1) | 131 (65.5) | 28 (59.0) | - | 65 (64.4) | 101 (62.5) | 51 (60.9) | 115 (64.4) | 109 (59.6) | 37 (78.4) | 19 (61.8) |
| Less physically active | 92 (71.6) | 76 (57.0) | 5 (39.7) | 139 (69.5) | 23.5 (48.9) | - | 60 (59.3) | 108 (67.2) | 50 (60.0) | 118 (66.1) | 118 (64.3) | 30 (63.2) | 20 (64.8) |
| Less time with friends | 91 (71.1) | 89 (66.2) | 6 (51.0) | 148 (73.7) | 25 (52.4) | - | 72 (71.6) | 107 (66.7) | 57 (68.0) | 123 (68.8) | 118 (64.3) | 39 (82.6) | 22 (72.3) |
| Finding school harder | 61 (47.7) | 56 (42.2) | 4 (32.0) | 91 (45.5) | 22 (46.7) | - | 45 (44.7) | 72 (45.0) | 36 (43.3) | 81 (45.6) | 79 (43.2) | 28 (59.3) | 10 (32.7) |
| **<u>Financial impacts</u>** |  |  |  |  |  |  |  |  |  |  |  |  |  |
| Employment status changed | 139 (40.5) | 134 (36.8) | 76 (51.8) | 134 (45.5) | 58 (30.0) | 5 (6.3) | 101 (35.7) | 172 (41.3) | 70 (31.4) | 202 (41.9) | 182 (43.1) | 62 (39.9) | 30 (22.6) |
| Worried about financial problems | 128 (37.1) | 161 (44.2) | 65 (44.1) | 135 (45.9) | 76 (39.5) | 12 (16.6) | 104 (35.8) | 184 (44.3) | 288 (40.8) | 201 (41.6) | 166 (39.5) | 73 (47.0) | 49 (37.4) |
| Unable to meet weekly expenses | 80 (23.2) | 75 (20.8) | 32 (21.9) | 64 (21.8) | 47 (24.6) | 11 (15.7) | 64 (21.9) | 91 (22.0) | 49 (21.7) | 107 (22.1) | 93 (22.0) | 30 (19.4) | 33 (24.8) |
|  | **M (95% CI)** | **M (95% CI)** | **M (95% CI)** | **M (95% CI)** | **M (95% CI)** | **M (95% CI)** | **M (95% CI)** | **M (95% CI)** | **M (95% CI)** | **M (95% CI)** | **M (95% CI)** | **M (95% CI)** | **M (95% CI)** |
| **<u>Mean negative impact on children<sup>#</sup></u>** | 3.6 (3.2, 4.0) | 3.5 (3.3, 3.7) | 2.9 (2.1, 3.8) | 3.7 (3.5, 3.8) | 3.2 (2.5, 3.9) | - | 3.5 (3.1, 3.9) | 3.6 (3.3, 3.8) | 3.4 (3.0, 3.9) | 3.6 (3.4, 3.8) | 3.5 (3.2, 3.7) | 3.8 (3.6, 4.0) | 3.4 (2.7, 4.1) |
| **<u>Mean financial burden<sup>†</sup></u>** | 2.8 (2.7, 3.0) | 2.9 (2.8, 3.0) | 2.8 (2.6, 3.1) | 2.9 (2.8, 3.0) | 2.9 (2.8, 3.1) | 2.4 (2.3, 2.6) | 2.8 (2.7, 3.0) | 2.9 (2.8, 3.0) | 2.8 (2.7, 3.0) | 2.9 (2.8, 3.0) | 2.8 (2.7, 2.9) | 3.0 (2.8, 3.2) | 2.9 (2.7, 3.0) |

1 respondent indicated 'other/prefer not to say' and is not included in weighted analyses presented in this table
\*Health conditions assessed included respiratory disease, asthma, chronic obstructive pulmonary disease, high blood pressure, cancer, heart disease, stroke, diabetes, depression and anxiety
\*\*Total number of participants that responded to the question regarding the impacts of COVID-19 on their relationship with their partner = 399; Total number of participants reporting having children = 262. Impacts on children are not reported for age group 70+ due to small numbers
Composite score comprising impact on screen time, physical activity, time with friends and schooling. Scale range: 1-5. Higher scores indicate more negative impact.
Scale range: 1-5. Higher scores indicate greater perceived financial burden.

Overall, 22.3% of participants reported feeling alone or lonely most or all of the time. The oldest age group (70 years or more) had the highest proportion of participants reporting feeling lonely or alone (45.5%, n=33) while those aged 30-49 years had the lowest proportion (17.6%; n=52). Similar proportions of males and females felt alone or lonely most or all of the time (21.8% and 22.8% respectively). 27.8% (n=81) of participants with inadequate health literacy reported feeling alone or lonely most or all of the time; this proportion was 18.5% for participants with adequate health literacy (n=77). In regards to language groups, the range was from 5.6% (n=2) for Hindi speakers to 51.2% (n=32) for Khmer speakers. See Tables 3 and 4. In the multivariable regression model, having two or more chronic illnesses (p<0.001) and university education (p<0.001) remained as significant correlates of feeling lonely or alone, with statistically significant differences also observed between language groups (p<0.001).

**Table 4.** Psychological, social and financial impacts, by language group (n=707)*

|  | Arabic | Assyria<br>n | Chinese | Croatia<br>n | Dari | Dinka | Hindi | Khmer | Samoan<br>/<br>Tongan | Spanish | All |
| --- | --- | --- | --- | --- | --- | --- | --- | --- | --- | --- | --- |
| **Variable** | **n (%)** | **n (%)** | **n (%)** | **n (%)** | **n (%)** | **n (%)** | **n (%)** | **n (%)** | **n (%)** | **n (%)** | **n (%)** |
| **<u>Psychological impacts</u>** |  |  |  |  |  |  |  |  |  |  |  |
| Nervous or stressed | 14 (17.9) | 22 (16.9) | 5 (6.0) | 40 (33.4) | 14 (31.9) | 24 (38.0) | 6 (13.4) | 36 (57.1) | 12 (29.0) | 5 (11.9) | 179 (25.3) |
| Alone or lonely | 19 (23.5) | 13 (9.5) | 5 (6.1) | 50 (41.1) | 8 (17.8) | 15 (24.4) | 2 (5.6) | 32 (51.2) | 8 (19.1) | 6 (14.2) | 158 (22.3) |
| **<u>Social impacts\*\*</u>** |  |  |  |  |  |  |  |  |  |  |  |
| Negative impact on relationship | 3 (6.7) | 2 (2.7) | 8 (20.1) | 40 (38.2) | 10 (32.9) | 6 (18.9) | 5 (14.7) | 8 (45.3) | 0 (0.0) | 5 (31.4) | 102 (25.5) |
| More time looking after children | 12 (69.8) | 40 (80.4) | 25 (87.0) | 33 (93.8) | 17 (83.7) | 25 (66.3) | 7 (36.7) | 16 (90.2) | 13 (72.6) | 4 (20.9) | 191 (72.8) |
| More screen time | 13 (73.6) | 30 (60.6) | 23 (81.0) | 34 (97.3) | 8 (39.2) | 24 (64.7) | 9 (49.4) | 10 (56.3) | 14 (81.0) | 1 (3.5) | 166 (63.3) |
| Less physically active | 15 (87.2) | 22 (44.8) | 25 (88.9) | 32 (91.4) | 6 (28.4) | 24 (63.3) | 11 (60.9) | 15 (85.3) | 13 (76.2) | 5 (23.1) | 168 (64.2) |
| Less time with friends | 8 (49.4) | 28 (55.5) | 25 (87.0) | 34 (97.3) | 14 (66.9) | 27 (72.9) | 8 (44.9) | 17 (100.0) | 14 (79.8) | 4 (20.9) | 180 (68.6) |
| Finding school harder | 12 (72.5) | 21 (42.5) | 9 (32.3) | 22 (63.4) | 3 (13.4) | 18 (49.5) | 4 (21.8) | 17 (77.5) | 14 (79.8) | - | 118 (44.9) |
| **<u>Financial impacts</u>** |  |  |  |  |  |  |  |  |  |  |  |
| Employment status changed | 29 (36.1) | 24 (18.2) | 30 (39.8) | 51 (41.9) | 25 (56.7) | 25 (40.3) | 20 (48.0) | 38 (59.7) | 18 (42.8) | 13 (29.7) | 272.8 (38.6) |
| Worried about financial problems | 19 (23.7) | 39 (29.0) | 14 (18.4) | 57 (46.8) | 26 (59.5) | 39 (62.3) | 17 (40.4) | 45 (71.3) | 23 (56.3) | 9 (21.4) | 288 (40.8) |
| Unable to meet weekly expenses | 12 (14.8) | 39 (28.4) | 21 (27.3) | 5 (3.9) | 8 (18.7) | 15 (24.5) | 3 (7.4) | 30 (47.7) | 15 (36.9) | 6 (14.9) | 155 (21.9) |
|  | **M (95% CI)** | **M (95% CI)** | **M (95% CI)** | **M (95% CI)** | **M (95% CI)** | **M (95% CI)** | **M (95% CI)** | **M (95% CI)** | **M (95% CI)** | **M (95% CI)** | **M (95% CI)** |
| **<u>Mean negative impact on children\*\*\*</u>** | 3.6 (3.2, 4.0) | 3.4 (3.1, 3.6) | 4.1 (3.8, 4.3) | 4.3 (4.1, 4.5) | 2.9 (2.3, 3.4) | 3.7 (3.3, 4.1) | 3.1 (2.6, 3.5) | 4.1 (3.9, 4.3) | 4.1 (3.7, 4.5) | 1.8 (0.7, 2.9) | 3.5 (3.3, 3.7) |
| **<u>Mean financial burden†</u>** | 2.7 (2.5, 3.0) | 2.7 (2.6, 2.9) | 2.6 (2.3, 2.9) | 2.7 (2.6, 2.9) | 3.4 (3.2, 3.6) | 3.1 (2.8, 3.3) | 2.8 (2.6, 3.0) | 3.6 (3.4, 3.8) | 3.0 (2.6, 3.5) | 2.1 (1.7, 2.6) | 2.9 (2.8, 2.9) |

\*1 respondent indicated 'other/prefer not to say' and is not included in weighted analyses presented in this table
\*\*Total number of participants that responded to the question regarding the impacts of COVID-19 on their relationship with their partner = 399; Total number of participants reporting having children = 262.
\*\*\*Composite score comprising impact on screen time, physical activity, time with friends and schooling. Scale range: 1-5. Higher scores indicate more negative impact
†Scale range: 1-5. Higher scores indicate greater perceived financial burden.

### Social impacts

Of the 399 participants who responded to the question regarding impacts of COVID-19 on their relationship with their partner, one quarter (25.5%) reported negative effects; 62.9% said that the pandemic had no effect and 11.7% said that it had had positive effects. We observed significant differences in reporting of negative impacts on relationships across language groups (p<0.001) and across age groups such that those aged <30 years had a significantly higher proportion of people reporting negative impacts compared to each other age group (30-49: p<0.001; 50-69: p<0.001; 70 and above: p=0.02). Those in the most disadvantaged IRSAD quintile reported more negative impacts compared to those in higher quintiles (p<0.01). We also observed significant differences in reporting of negative impacts on relationships based on financial burden (p<0.001) and psychological variables (alone/lonely - p<0.001; nervous/stressed - p<0.001).

Of the two hundred and sixty-two participants who reported having children aged less than 18 years, 72.8% reported spending more time looking after their children as a result of the pandemic (n=191). The majority agreed (somewhat or strongly) that COVID-19 has meant that their children spent less time with friends (68.5%), are less physically active (64.2%), and have more screen time (63.3%). Across the entire sample, 44.9% agreed that their children were finding school harder. Mean perceived negative impact on children was rated 3.5 (out of 5; 95% CI= 3.3 to 3.7). Reporting of negative impacts on children was significantly associated with the most disadvantaged IRSAD quintile (p=0.02) and with chronic illness, with participants with one (p=0.01) or two or more (p<0.001) chronic illnesses significantly more likely to report negative impacts compared to those without chronic illness. Reporting of negative impacts on children also varied significantly across language groups (p<0.001). See Supplementary Table 2.

### Financial impacts

Overall, 38.6% of participants reported that their employment status has changed because of COVID-19. This was most commonly a reduction in hours of employment. See Figure 1. In total, 63.3% of participants reported somewhat or more worry about financial problems as a result of the COVID-19 pandemic, and 53.7% reported that they were having difficulty meeting their financial expenses.

Mean perceived financial burden was 2.9 on a five-point scale (95% Confidence Interval [CI]=2.8 to 2.9). Perceived financial burden was similar across genders, health literacy categories and age groups with the exception of the oldest age group (70+ years) which had a lower mean financial burden score of 2.4 (95% CI= 2.3 to 2.6). In the multivariable regression model, mean perceived financial burden was significantly lower for the oldest age group compared to the youngest after controlling for other sociodemographic factors (p<0.001).

As well as differences by age, we also observed significant differences in mean perceived financial burden across language groups (p<0.001) (Supplementary Table 3). People with one chronic illness (p=0.01) or two or more (p<0.001) reported significantly more financial burden compared to those without chronic illness.

## DISCUSSION

This is the largest Australian survey exploring the impacts of COVID-19 among people who primarily speak a language other than English. Even prior to the July 2021 COVID-19 outbreak in New South Wales, which has disproportionately impacted the communities and geographical areas included in this study, we observed broad negative psychological, social and financial impacts of the pandemic. Over one quarter of the sample reported feeling nervous or stressed most or all of the time, and twenty-two percent felt lonely or alone most or all of the time. Over half worried about financial problems and reported being somewhat or less able to meet their weekly expenses. One quarter of participants reported negative impacts on their spousal relationship and the majority of participants with children under 18 years reported that even out of lockdown their children spent less time with friends as a result of the pandemic (68.5%), were less physically active (64.2%) and had more screen time (63.3%). Regression analyses consistently showed distinct patterns of COVID-19 impacts for different language groups and more negative outcomes for those living with chronic illness and comorbidities.

The impacts of COVID-19 have been explored across a number of countries with different population groups. Direct comparisons are difficult on account of varying survey items, different data collection timepoints, and wide-ranging case numbers, morbidity and mortality from COVID-19 worldwide. However, psychological impacts found in this study are comparable to our national survey conducted in April 2020, at the outset of the pandemic when stay at home orders had been in place for 3 weeks. In this earlier study, we found that 26% of participants reported feeling nervous or stressed most or all of the time, and 27% percent felt lonely or alone most or all of the time (15). Nationally-representative data from the Australian Bureau of Statistics similarly showed that in June 2021, one in five (20%) Australians experienced high or very high levels of psychological distress in the last four weeks, and 28% of people 18 years and over reported feeling nervous in that survey (10). Previous work has also confirmed negative impacts of COVID on children’s social connectedness and amount of screen time (25, 26). This is the first time to demonstrate these impacts among a large sample of people who speak a language other than English at home.

### Implications

A multi-level, whole-of-government approach to address the impacts of COVID-19 for culturally and linguistically-diverse communities. This must necessarily involve a host of coherent multisectoral actions. Policy and sustainable infrastructure is needed to ensure the readiness of the system to map and meet evolving needs of a multicultural society and support meaningful engagement of communities to co-design innovative, tailored and culturally-safe support packages (27). Qualitative studies have highlighted a large number of community-driven initiatives and actions that have emerged as a response to COVID-19, as well as embodied and communal ways of coping (28). Using a strengths-based perspective, we must acknowledge the multiple capacities and resources of our culturally and linguistically diverse communities and provide properly-resourced opportunities to work directly with them to address unique challenges that they face. Timely, understandable and culturally-appropriate information about financial, social and mental health resources and services must be prioritised in line with Lancet Migration’s call for responsible, transparent and migrant-inclusive public information strategies (29).

### Strengths and limitations

This study was co-designed by researchers and multicultural health service staff, and enabled through recruitment methods that are inclusive and reduce barriers to participation, such as translated versions of the survey, engagement of interpreters and multicultural health staff who are trusted in their communities, and use of multiple recruitment methods (including through community events and networks). This approach wholly aligns with the Framework of Culturally Competent Health Research (16). However, practical constraints limited the number of languages we could include, and restricted data collection to three regions in Greater Sydney only. We also used convenience sampling methods.

To reduce survey length and burden on participants we purposefully selected a small number of items from validated measures or our previous research to explore psychological, social and financial impacts, or co-designed them specifically for this study. Self-report may have introduced recall and social desirability bias.

Finally, the results of this study reflect a particular point in time when there were very low numbers of community-acquired cases of COVID-19 in Australia, and for the most part, no government-imposed restrictions on movement and activities in New South Wales. It is likely that psychological wellbeing outcomes and financial and social stress have worsened since the July 2021 outbreak and the imposition of stay-at-home orders, in line with previous research (25, 30). We are unable to explore changes in impacts over time in this study.

## Conclusion

Culturally and linguistically diverse communities experience significant impacts of COVID-19, with distinct patterns of impacts for different language groups. We must work with communities to address unique challenges they face and tailor interventions and supports accordingly. As COVID-19 continues to disproportionately impact the most culturally and linguistically diverse communities in Sydney and worldwide, responses must too reflect the diversity of our communities through co-production and tailored support packages.

## Data Availability

Deidentified participant data are available from the first author (ORCID identifier: 0000-0001-6106-6298) upon reasonable request.

## FIGURE LEGENDS

Figure 1. Change in employment

## AUTHORS’ CONTRIBUTIONS

**Muscat DM** - Formal analysis, Investigation; Data Curation; Writing - Original Draft; **Ayre J** - Conceptualization, Methodology, Formal analysis, Investigation; Data Curation; Writing - Review & Editing; Project administration; **Mac O** - Formal analysis, Investigation; Data Curation; Writing - Review & Editing; Project administration; **Batcup C** - Conceptualization, Methodology, Investigation; Writing - Review & Editing; Project administration; **Cvejic E** - Formal analysis, Writing - Review & Editing Pickles K; Conceptualization, Methodology, Writing - Review & Editing; **Dolan H** - Conceptualization, Methodology, Writing - Review & Editing; **Bonner C** - Conceptualization, Methodology, Writing - Review & Editing; **Mouwad D** - Conceptualization, Methodology, Investigation; Writing - Review & Editing; **Zachariah D** - Conceptualization, Methodology, Investigation; Writing - Review & Editing; **Turalic U** - Conceptualization, Methodology, Investigation; Writing - Review & Editing; **Santalucia Y** - Conceptualization, Methodology, Investigation; Writing - Review & Editing; **Chen T** - Conceptualization, Methodology, Investigation; Writing - Review & Editing **Vasic G** - Conceptualization, Methodology, Investigation; Writing - Review & Editing; **McCaffery KJ** - Conceptualization, Methodology, Formal analysis, Writing - Review & Editing; Project administration.

## ETHICS STATEMENT

This study was approved by Western Sydney Local Health District Human Research Ethics Committee (Project number 2020/ETH03085). All participants provided informed consent to participate. This manuscript does not contain any personal or medical information about an identifiable individual.

