## Supplemental Table for "Psychological, social and financial impact of COVID-19 on culturally and linguistically diverse communities: a cross-sectional Australian study"

**Supplementary Table 1. Multiple regression model of factors associated with negative psychological impacts (n=707)***

| **Predictor** | **Nervous/Stressed** | | | | **Alone/Lonely** | | | |
| --- | --- | --- | --- | --- | --- | --- | --- | --- |
|  | **Unadjusted analysis** | | **Adjusted analysis** | | **Unadjusted analysis** | | **Adjusted analysis** | |
|  | **OR (95% CI)** | **P value** | **OR (95% CI)** | **P value** | **OR (95% CI)** | **P value** | **OR (95% CI)** | **P value** |
| **Gender*** |  |  |  |  |  |  |  |  |
| Male | Reference |  | Reference |  | Reference |  | Reference |  |
| Female | 1.51 (1.03 to 2.22) | 0.03 | 1.59 (1.03 to 2.45) | 0.04 | 1.05 (0.70 to 1.60) | 0.80 | 1.01 (0.65 to 1.57) | 0.97 |
| **Age group** |  | **0.06** |  | **0.65** |  | **<0.001** |  | **0.36** |
| 18-29 | Reference |  | Reference |  | Reference |  | Reference |  |
| 30-49 | 1.07 (0.59 to 1.95) | 0.82 | 0.76 (0.39 to 1.48) | 0.42 | 0.84 (0.42 to 1.70) | 0.63 | 1.54 (0.78 to 3.06) | 0.22 |
| 50-69 | 1.62 (0.89 to 2.95) | 0.12 | 0.99 (0.47 to 2.11) | 0.99 | 1.16 (0.57 to 2.33) | 0.68 | 1.41 (0.62 to 3.23) | 0.42 |
| >70 | 2.04 (1.00 to 4.15) | 0.05 | 1.09 (0.41 to 2.88) | 0.87 | 3.30 (1.53 to 7.12) | <0.001 | 0.93 (0.35 to 2.48) | 0.88 |
| **Comorbidity**** |  | **<0.001** |  | **0.01** |  | **<0.001** |  | **<0.001** |
| 0 | Reference |  | Reference |  | Reference | <0.001 | Reference |  |
| 1 | 0.60 (0.39 to 0.94) | 0.03 | 1.34 (0.77 to 2.32) | 0.30 | 1.48 (0.90 to 2.44) | 0.12 | 0.80 (0.45 to 1.44) | 0.460 |
| 2+ | 0.44 (0.27 to 0.70) | <0.001 | 2.39 (1.35 to 4.24) | <0.001 | 2.74 (1.67 to 4.51) | <0.001 | 0.34 (0.18 to 0.64) | <0.001 |
| **Lowest ISRAD quintile** | 1.16 (0.77 to 1.74) | 0.47 | 1.41 (0.86 to 2.31) | 0.17 | 0.80 (0.51 to 1.24) | 0.32 | 1.08 (0.64 to 1.84) | 0.77 |
| **University education** | 0.47 (0.30 to 0.73) | <0.001 | 1.28 (0.71 to 2.32) | 0.41 | 0.43 (0.26 to 0.74) | <0.001 | 1.10 (0.58 to 2.08) | <0.001 |
| **Adequate health literacy** | 0.62 (0.42 to 0.90) | 0.01 | 0.68 (0.39 to 1.19) | 0.18 | 0.59 (0.39 to 0.9) | 0.01 | 1.17 (0.67 to 2.04) | 0.57 |
| **English-language proficiency** | 0.54 (0.37 to 0.78) | <0.001 | 0.88 (0.50 to 1.57) | 0.68 | 0.49 (0.33 to 0.74) | <0.001 | 0.93 (0.51 to 1.72) | 0.83 |
| **Years living in Australia** | | **0.12** |  | **0.70** |  | **0.42** |  | **0.870** |
| 5 years or less | Reference |  | Reference |  | Reference |  | Reference |  |
| 6 to 10 years | 1.36 (0.70 to 2.64) | 0.36 | 1.22 (0.58 to 2.53) | 0.60 | 1.26 (0.58 to 2.72) | 0.56 | 0.88 (0.42 to 1.84) | 0.73 |
| More than 10 years | 1.27 (0.74 to 2.18) | 0.38 | 1.19 (0.61 to 2.34) | 0.61 | 1.22 (0.63 to 2.37) | 0.56 | 1 (0.51 to 1.95) | 0.99 |
| Born in Australia | 0.51 (0.21 to 1.26) | 0.14 | 0.73 (0.25 to 2.18) | 0.58 | 0.60 (0.21 to 1.70) | 0.34 | 1.38 (0.43 to 4.39) | 0.59 |
| **Language group***** | - | **<0.001** | **-** | **<0.001** | **-** | **<0.001** | **-** | **<0.001** |
| **Perceived public health threat** | 1.15 (1.08 to 1.22) | <0.001 | 1.08 (0.99 to 1.18) | 0.07 | 1.12 (1.05 to 1.20) | <0.001 | 0.93 (0.85 to 1.03) | 0.15 |
| **Mean financial burden** | 1.96 (1.55 to 2.48) | <0.001 | 1.82 (1.42 to 2.33) | <0.001 | - | - | - | - |

NB: All regression models also control for date of survey completion (binary variable, before/after 23 June when restrictions in Greater Sydney were imposed).

*1 respondent indicated ‘other/prefer not to say’ and is not included in weighted analysis

**Health conditions assessed included respiratory disease, asthma, chronic obstructive pulmonary disease, high blood pressure, cancer, heart disease, stroke, diabetes, depression and anxiety

***Individual comparisons for language group not presented as there is no specific contrast that is pragmatically relevant . Khmer was selected as the reference language group as this subsample was of adequate size (n>50) and had the highest proportion of people reporting negative psychological impacts.

**Supplementary Table 2. Multiple regression model of factors associated with negative social impacts**

|  | **Negative impact on relationships (n=399)*** | | | | **Negative impact on children (n=262)**** | | | |
| --- | --- | --- | --- | --- | --- | --- | --- | --- |
| **Predictor** | **Unadjusted analysis** | | **Adjusted analysis** | | **Unadjusted analysis** | | **Adjusted analysis** | |
|  | **OR (95% CI)** | **P value** | **OR (95% CI)** | **P value** | **B(95%CI)** | **P value** | **B (95% CI)** | **P value** |
| **Gender** |  |  |  |  |  |  |  |  |
| Male | Reference |  | Reference |  | Reference |  | Reference |  |
| Female | 1.16 (0.65 to 2.09) | 0.62 | 1.21 (0.72 to 2.03) | 0.47 | -0.12 (-0.54 to 0.31) | 0.59 | -0.17 (-0.41 to 0.08) | 0.18 |
| **Age group** |  | **0.15** |  | **0.03** |  | **0.13** |  | **0.12** |
| 18-29 | Reference |  | Reference |  | Reference |  | Reference |  |
| 30-49 | 0.32 (0.09 to 1.09) | 0.07 | 0.27 (0.11 to 0.65) | <0.001 | 0.75 (-0.13 to 1.62) | 0.10 | 0.70 (-0.03 to 1.43) | 0.06 |
| 50-69 | 0.57 (0.13 to 2.41) | 0.44 | 0.41 (0.17 to 0.98) | <0.001 | 0.32 (-0.78 to 1.41) | 0.57 | 0.46 (-0.35 to 1.27) | 0.26 |
| >70 | 0.40 (0.07 to 2.30) | 0.30 | 0.30 (0.11 to 0.86) | 0.02 | 0 (-1.32 to 1.33) | 1.00 | 0.44 (-1.40 to 2.28) | 0.64 |
| **Chronic illness***** |  | **0.63** |  | **0.70** |  | **0.15** |  | **<0.001** |
| 0 | Reference |  | Reference |  | Reference |  | Reference |  |
| 1 | 0.81 (0.39 to 1.68) | 0.57 | 1.11 (0.61 to 2.00) | 0.74 | 0.33 (-0.02 to 0.68) | 0.070 | 0.37 (0.09 to 0.65) | 0.01 |
| 2+ | 1.28 (0.50 to 3.24) | 0.60 | 1.33 (0.68 to 2.60) | 0.40 | -0.07 (-0.82 to 0.68) | 0.850 | 0.76 (0.27 to 1.26) | <0.001 |
| **Lowest IRSAD quintile** | 0.34 (0.14 to 0.82) | 0.02 | 0.34 (0.17 to 0.66) | <0.001 | -0.17 (-0.67 to 0.33) | 0.50 | 0.40 (0.07 to 0.72) | 0.02 |
| **University education** | 1.87 (0.68 to 5.13) | 0.23 | 0.50 (0.25 to 1.02) | 0.06 | -0.12 (-0.53 to 0.29) | 0.57 | -0.02 (-0.36 to 0.32) | 0.91 |
| **Adequate health literacy** | 0.41 (0.21 to 0.81) | 0.01 | 0.80 (0.48 to 1.35) | 0.41 | 0.08 (-0.38 to 0.53) | 0.75 | 0.22 (-0.08 to 0.53) | 0.15 |
| **English-language proficiency** | 1.46 (0.66 to 3.21) | 0.35 | 1.02 (0.61 to 1.71) | 0.95 | -0.31 (-0.64 to 0.02) | 0.06 | -0.11 (-0.41 to 0.19) | 0.46 |
| **Years living in Australia** |  | **0.13** |  | **0.53** |  | **0.91** |  | **0.99** |
| 5 years or less | Reference |  | Reference |  | Reference |  | Reference |  |
| 6 to 10 years | 1.14 (0.45 to 2.93) | 0.78 | 1.78 (0.73 to 4.34) | 0.21 | 0.04 (-0.64 to 0.71) | 0.91 | -0.09 (-0.57 to 0.39) | 0.72 |
| More than 10 years | 0.47 (0.18 to 1.20) | 0.12 | 1.01 (0.53 to 1.93) | 0.98 | -0.07 (-0.46 to 0.32) | 0.73 | -0.03 (-0.41 to 0.36) | 0.89 |
| Born in Australia | 0.41 (0.08 to 2.15) | 0.29 | 1.06 (0.34 to 3.35) | 0.92 | -0.34 (-1.4 to 0.72) | 0.52 | 0 (-0.59 to 0.59) | 1.00 |
| **Language group^#^** |  | **<0.001** |  | **<0.001** |  | **<0.001** |  | **<0.001** |
| **Perceived public health threat^†^** | 0.97 (0.85 to 1.10) | 0.59 | 1.06 (0.99 to 1.15) | 0.10 | - | - | - | - |
| **Mean financial burden^†^** | 1.70 (1.14 to 2.54) | 0.01 | 1.88 (1.38 to 2.56) | <0.001 | - | - | - | - |
| **Feeling lonely / alone^†^** | 0.98 (0.40 to 2.40) | 0.96 | 0.37 (0.21 to 0.64) | <0.001 | - | - | - | - |
| **Feeling nervous / stressed^†^** | 0.33 (0.14 to 0.77) | 0.01 | 0.29 (0.17 to 0.49) | <0.001 | - | - | - | - |

NB: All regression models also control for date of survey completion (binary variable, before/after 23 June when restrictions in Greater Sydney were imposed).

*Total number of participants that responded to the question regarding the impacts of COVID-19 on their relationship with their partner

**Total number of participants reporting having children

***Health conditions assessed included respiratory disease, asthma, chronic obstructive pulmonary disease, high blood pressure, cancer, heart disease, stroke, diabetes, depression and anxiety

**^#^**Individual comparisons for language group not presented as there is no specific contrast that is pragmatically relevant . Khmer was selected as the reference language group as this subsample was of adequate size (n>50) and had the highest proportion of people reporting negative impacts on relationships.

**^†^**Variable not included in the regression model of factors associated with negative impacts on children

**Supplementary Table 3. Multiple regression model of factors associated with financial burden (n=707)***

|  | **Unadjusted analysis** | | **Adjusted analysis** | |
| --- | --- | --- | --- | --- |
|  | **B (95% CI)** | **P value** | **B (95% CI)** | **P value** |
| **Gender** |  |  |  |  |
| Male | Reference |  | Reference |  |
| Female | 0.03 (-0.15 to 0.21) | 0.77 | 0.01 (-0.13 to 0.15) | 0.89 |
| **Age group** |  | **<0.001** |  | **<0.001** |
| 18-29 | Reference |  | Reference |  |
| 30-49 | 0.07 (-0.19 to 0.34) | 0.58 | 0.08 (-0.15 to 0.32) | 0.49 |
| 50-69 | 0.09 (-0.20 to 0.38) | 0.54 | 0.03 (-0.22 to 0.29) | 0.80 |
| >70 | -0.40 (-0.67 to -0.12) | 0.01 | -0.51 (-0.82 to -0.20) | <0.001 |
| **Comorbidity**** |  | **0.14** |  | **<0.001** |
| 0 | Reference |  | Reference |  |
| 1 | 0.21 (0 to 0.41) | 0.05 | 0.26 (0.06 to 0.46) | 0.01 |
| 2+ | 0.07 (-0.12 to 0.26) | 0.48 | 0.35 (0.15 to 0.54) | <0.001 |
| **Lowest IRSAD quintile** | -0.01 (-0.21 to 0.18) | 0.91 | -0.06 (-0.22 to 0.11) | 0.50 |
| **University education** | -0.27 (-0.46 to -0.09) | <0.001 | -0.18 (-0.36 to 0.01) | 0.06 |
| **Adequate health literacy** | 0.05 (-0.13 to 0.24) | 0.56 | 0.14 (-0.06 to 0.33) | 0.16 |
| **English-language proficiency** | -0.09 (-0.25 to 0.07) | 0.27 | -0.12 (-0.32 to 0.08) | 0.24 |
| **Years living in Australia** |  | **0.01** |  | **0.24** |
| 5 years or less | Reference |  | Reference |  |
| 6 to 10 years | 0.14 (-0.11 to 0.38) | 0.27 | 0.05 (-0.19 to 0.30) | 0.67 |
| More than 10 years | -0.17 (-0.37 to 0.02) | 0.07 | -0.12 (-0.32 to 0.09) | 0.26 |
| Born in Australia | -0.33 (-0.77 to 0.11) | 0.14 | -0.21 (-0.61 to 0.19) | 0.31 |
| **Language group***** | **-** | **<0.001** | **-** | **<0.001** |

NB: All regression models also control for date of survey completion (binary variable, before/after 23 June when restrictions in Greater Sydney were imposed).

*1 respondent indicated ‘other/prefer not to say’ and is not included in weighted analysis

**Health conditions assessed included respiratory disease, asthma, chronic obstructive pulmonary disease, high blood pressure, cancer, heart disease, stroke, diabetes, depression and anxiety

***Individual comparisons for language group not presented as there is no specific contrast that is pragmatically relevant . Khmer was selected as the reference language group as this subsample was of adequate size (n>50).
